# Symbolic regression analysis of interactions between first trimester maternal serum adipokines in pregnancies which develop pre-eclampsia

**DOI:** 10.1101/2022.06.29.22277072

**Authors:** Casper Wilstrup, Paula L. Hedley, Line Rode, Sophie Placing, Karen R. Wøjdemann, Anne-Cathrine Shalmi, Karin Sundberg, Michael Christiansen

## Abstract

**Objectives:** Pre-eclampsia (PE) is an important cause of perinatal morbidity and mortality. Despite an elusive pathophysiology, PE has been associated with changes in maternal serum concentrations of adipokines in early pregnancy. We hypothesized, that symbolic regression might identify interactions between adipokines and PE, which may have eluded regression and Bayesian models.

**Methods:** In this nested case-control sub-study, of the Copenhagen First Trimester Screening Study, data regarding maternal weight and serum concentrations of PAPP-A, leptin (Lp), soluble leptin receptor (sLR), adiponectin, and resistin (Re) was available from 423 first trimester pregnancies (gestational week 10^+3^– 13^+6^), 126 of which developed PE. Symbolic regression with QLattice/Feyn 2.1 was used to identify models comprising two-interactions between up-to three markers.

**Results:** The optimal mathematical model exhibited a non-linear relation between Re and the combined effect of sLR and Lp. The model was dependent, in a Gaussian way, on the level of Re. The receiver operating characteristic (ROC) curve of the model viz. identification of PE cases in first trimester had an AUC of 0.81 and a modelled DR of 40 % for a FPR of 4 %. Symbolic regression outperformed logistic regression of the same parameters with a ROC with AUC = 0.77, and a DR of 7 % for a 3 % FPR.

**Conclusions:** Symbolic regression identified non-linear interactions between Lp, sLR and Re concentrations in first trimester pregnancy serum of pregnancies which later developed PE. Non-linear interactions suggest new pathophysiological pathways and may help in designing more efficient screening protocols for PE.

## Introduction

Pre-eclampsia (PE) is a frequent (3-5 %) adverse outcome of pregnancy characterized by elevated blood pressure and proteinuria or other organ failure occurring from gestational week 20 [1]. The condition is an important contributor to maternal [2, 3] and perinatal — due to growth disturbance, placental abruption, and iatrogenic premature birth [4] — mortality and morbidity. The prevalence of PE is higher in low- and middle-income countries [5]. PE has been estimated to cause ∼100K maternal deaths annually, through the development of symptomatic hypertension, eclampsia, and multiple organ involvement [6, 7]. It is of significance for outcome that high-risk patients are identified, carefully monitored, and offered preventive aspirin treatment, early [4, 8, 9]. PE is also associated with an increased occurrence of metabolic syndrome in both mother and foetus, emphasizing the significance of preventive strategies [1].

The aetiology of PE remains elusive, but several lines of evidence suggest that PE may be caused by defects in early trophoblast differentiation [10, 11]. This, in turn, may lead to impaired placentation [12], impaired trophoblast transformation of the uterine spiral arteries and placental hypoperfusion, which could result in endothelial dysfunction and elicit the maternal syndrome. Most of these processes can be monitored through biochemical markers found in maternal serum that, together with PE-associated demographic or clinical markers [13, 14], may be combined to create risk algorithms, and such algorithms have been constructed by FMF, ACOG and NICE and a number of other organisations [15], and clinically implemented with variable efficiency [16]. A more extensive use of electronic patient records, better clinical phenotyping, and use of machine learning (ML) may improve clinical usefulness of PE screening [17, 18].

A plethora of first trimester biochemical markers for the development of PE have been identified [19, 20], e.g. placenta growth factor [21], ADAM12 [22], PAPP-A [23], resistin (Re) [24], PP13 [25] and free leptin-index (leptin (Lp)/ soluble leptin receptor (sLR) [25, 26]. Mean arterial blood pressure is a frequently suggested marker [15] and Doppler ultrasonography of the uterine artery – in the form of a pulsatility index - has also been reported as a first trimester marker for the subsequent development of PE [27]. However, despite decades of research and an increasing number of first trimester PE markers, PE screening performance is still poorer than first trimester Down syndrome screening [14, 16, 19]. The association between metabolic syndrome in the mother and PE makes it relevant to assess the role of biomarkers reflecting maternal metabolic status, e.g. maternal weight, leptin, adiponectin, resistin, PAPP-A, in first trimester screening for PE.

All attempts to identify combinations of markers have used variations of linear, log-linear or Gaussian combinations of markers [15, 28]. Frequently based on assumptions about the distribution of markers as a function of gestational age and basic clinical information [14, 16, 19]. Machine learning (ML) models, with their promise of integrating multiple “omics” layers [29] and, potential to revolutionise individual patient treatment [30], have recently been introduced, albeit not yet in diagnostic use [31]. ML procedures are frequently of a “black-box” type, not revealing any underlying organization of parameters allowing for interpretability [32]. However, symbolic regression, exemplified by the QLattice method, works well with small datasets [33, 34], and returns a series of signal paths that are ordered by information content [35].

These signal paths reflect the interaction between the involved parameters [32], thus allowing for pathophysiologically relevant interpretation as well as construction of more efficient screening algorithms. This “white-box” characteristic makes symbolic regression an excellent exploratory research tool [18]. Thus, symbolic regression can be used to solve problems where the mathematical form of the data-generating process cannot be assumed a priori. This is in contrast with the typical regression problem where parameters are fitted to a fixed mathematical model, such as generalized linear models or logistic regression.

We hypothesized that a major reason for the inability to construct satisfactory risk algorithms is a lack of understanding of the complicated interactions between biochemical and clinical markers. Thus, the construction of statistical models based on generalized linear models may overlook the information content to be found in non-linear interactions between markers. In order to test this hypothesis we:

1. Examined whether previously reported metabolic maternal serum markers could be combined into a discriminatory mathematical function using symbolic regression.
2. Tested the screening performance of a symbolic regression based discriminatory function as compared to a log-linear discrimination.

## Materials and Methods

A nested case-control sub-study of the Copenhagen First Trimester Screening Study [36] was performed. Previously, several sub-studies of prenatal screening markers have been published [24-26, 37-40]. Out of 6,441 pregnant women for which serum samples were available; 160 verified PE pregnancies with proteinuria (2.5 %) were identified. Patients that developed PE were identified from the Danish National Patient Register [41, 42], and their files were retrieved and examined carefully for compliance with diagnostic criteria. Furthermore, to participate in the current study all patients had to have all the examined clinical parameters registered, and serum had to be available for the biochemical analysis. One hundred and twenty six PE pregnancies fulfilled these criteria [22]. Controls were selected at random but matched for maternal age, number of previous pregnancies and gestational age at biochemical examination. Gestational age was determined by crown-rump-length (CRL). Gestational age at birth was calculated using CRL and the date of birth [43]. All samples were collected in dry containers and kept at 4° C for a maximum of 48 hours until delivery to Statens Serum Institut. Samples were subsequently stored at – 20° C until analysis. Clinical and demographic information about patients and controls are given in Table 1. The biomarkers PAPP-A, resistin (Re), leptin (Lp), adiponectin and leptin soluble receptor (sLR) were quantitated in maternal serum as previously described [44-48].

**Table 1.** Demographic and clinical and paraclinical parameters of PE cases and controls.

| **Parameter** | **PE (n=126)** | **Controls (n=297)** | ***p-value*** |
| --- | --- | --- | --- |
| **Maternal age (years), mean (±SD)** | 30.6 (±4.56) | 30.3 (±4.21) | 0.631 |
| **Maternal weight (kg), median (IQR)** | 68.0 (62.0-75.0) | 62.0 (57.0-70.0)* | <0.0002 |
| **Maternal BMI (kg/m<sup>2</sup>), median (IQR)** | 24.0 (21.7-26.4) | 21.7 (20.1-24.0)* | <0.001 |
| **Primigravida, n (%)** | 74 (58.7) | 165 (55.6) | 0.75 |
| **Primipara, n (%)** | 101 (80.2) | 242 (81.5) | 0.79 |
| **Smoking, n (%)** | 16 (12.7) | 41 (13.8) | 0.72 |
| **Spontaneous conception, n (%)** | 120 (95.2) | 278 (93.6) | 0.29 |
| **Spontaneous labor onset, n (%)** | 32 (25.4) | 241 (81.1)* | <0.001 |
| **Crown rump length (mm), median (IQR)** | 67.0 (61.0-72.0) | 67.0 (61.0-74.0) | 0.85 |
| **Nuchal translucency (mm), median (IQR)** | 1.6 (1.3-1.9) | 1.5 (1.3-1.9) | 0.51 |
| **GA at birth (days), median (IQR)** | 275.5 (264.0-283.0) | 284.0 (278.0-290.0)* | 4.49e-06 |
| **GA at test (days), median (IQR)** | 90.5 (87.0 – 93.0) | 90.0 (88.0 – 94.0) | 1 |
| **GA below 34+0 weeks, n (%)** | 10 (7.9) | 0 (0.0)* | <0.001 |
| **PAPP-A (mIU/L), median (IQR)** | 3064 (1952 - 4576) | 3874 (2605 - 5938) | 0.028 |
| **hCG (IU/L), median (IQR)** | 42.90 (29.60 – 58.90) | 41.75 (26.95 – 63.10) | 0.975 |
| **Leptin (ug/L), median (range)** | 22.64 (5.00 – 70.14) | 14.1 (2.0 – 54.2)* | 1.47e-06 |
| **Leptin sR (ug/L), median (range)** | 27.8 (14.3 – 100.9) | 36.9 (15.1 – 97.5)* | 2.04e-13 |
| **Adiponectin (mg/L), median (range)** | 3.7 (1.6 – 10.2) | 4.5 (1.4 – 17.3)* | 7.58e-06 |
| **Resistin (ug/L), median (range)** | 21.89 (6.23 – 58.03) | 24.38 (4.94 – 82.96) | 0.177 |

As clinical information was collected in 1997 – 2001, we used the pre-2014 diagnostic criteria of PE of The International Society for the Study of Hypertension in Pregnancy (ISSHP) criteria to define PE [49]. Thus, the diagnosis of PE required hypertension, defined as either a systolic blood pressure > 140 mm Hg or a diastolic blood pressure > 90 mm Hg occurring in a previously normotensive woman after 20 weeks of gestation in combination with proteinuria (> 0.3 g per 24 hours or dipstick urine analysis >1+). In the current study we did not differentiate between mild and severe PE, early and late PE, or PE with and without IUGR.

### Statistical analysis

The analyses were performed using the QLattice/Feyn v 2.1 (Abzu, Copenhagen, Denmark) [35]. Briefly, the QLattice produces a collection of potential mathematical functions selected based on a categorical cross-entropy loss function defined by the discrimination between controls and PE pregnancies. Symbolic regression was performed with the restriction that the maximum number of markers and functions was five. The markers included in the analysis were adiponectin, PAPP-A, Lp, Re, sLR and maternal weight. Logistic regression and receiver operating characteristic (ROC) curve creation are features of the QLattice/Feyn program and confidence intervals of modelled screening performance were established with the quantile bootstrap method [50], commonly known as reverse percentile bootstrapping.

### Ethics

Blood sampling was performed as part of the Copenhagen First Trimester Screening Study and approved by the Scientific Ethics Committee for Copenhagen and Frederiksberg Counties, (No.(KF) 01-288/97) and The Data Protection Agency [36].

## Results

The symbolic regression identified a model comprising the parameters Lp, sLR, and Re as the model giving the optimal discrimination between PE and controls, neither adiponectin, maternal weight nor PAPP-A improved the discriminatory ability. The model (Figure 1) comprises a Gaussian interaction between Re and Lp followed by an addition of sLR. The discriminatory function is given in Figure 1B. The Gaussian interaction between Re and Lp followed by an addition of sLR is reflected in the nomogram of Re and sLR versus the probability of PE (Figure 2A), where it is clear that increasing sLR decreases the risk of PE, but that the iso-risk level depends on the Re level (see below). The partial derivative risk curves, figures 2B – 2D, demonstrate the Gaussian relation between Re, Lp and risk for PE.

**Figure 1.**
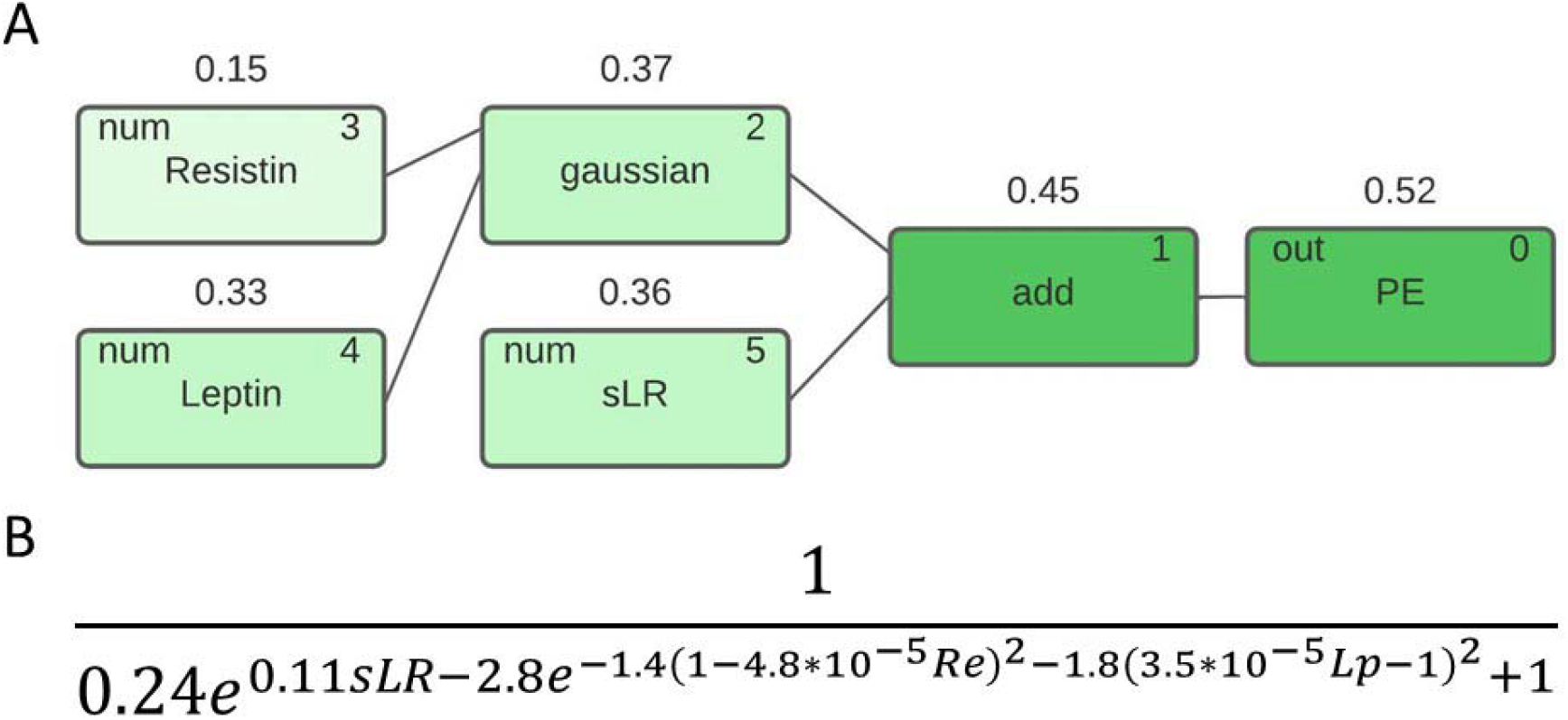
**A**. Model signal path in the symbolic regression model for PE. The numbers above the nodes are the Pearson correlation coefficient with the target after each mathematical operation. **B**. The mathematical equation describing the discriminatory function.

**Figure 2.**
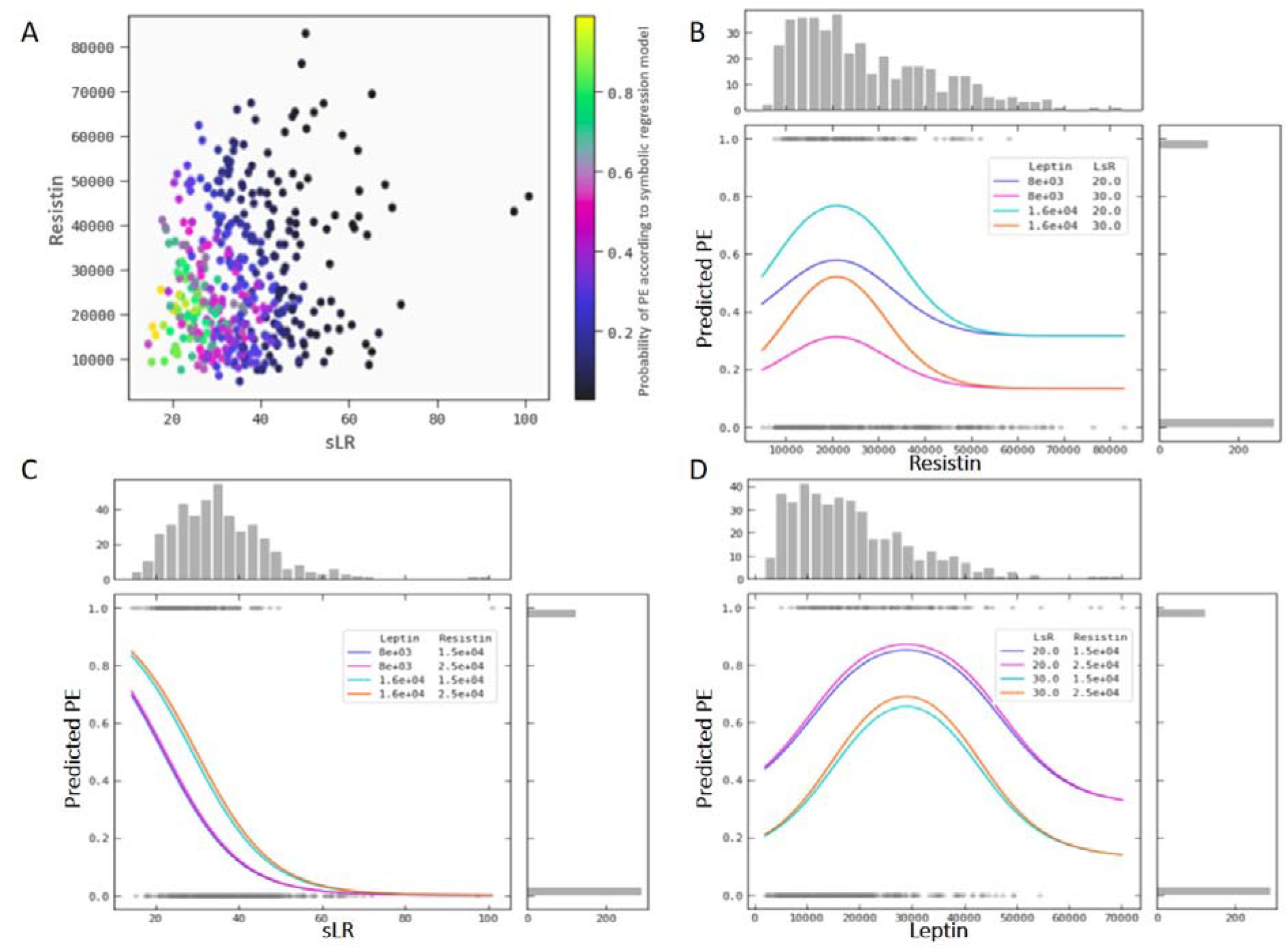
**A**. Nomogram depicting the risk of PE as a function of the combined Re and sLR level. **B**. Risk of PE as a function of Re level for different levels of Lp and sLR. **C**. Risk of PE as a function of sLR level for different levels of Re and Lp. **D**. Risk of PE as a function of Lp level for different levels of sLR and Re.

The symbolic regression model, with an AUC of 0.81, outperformed the equivalent logistic regression model (involving the same parameters) with an AUC of 0.77 (Figure 3). Using an estimated prevalence of 4 % for PE in the Danish population we modelled the expected performance of screening strategies for PE using the two models (Table 2) and found that the symbolic regression model offers a DR of 40 % for a FPR of 4 % as compared to a DR of 7 % for a FPR of 3 % for the logistic regression model. Indeed, the symbolic regression model out performs the logistic regression particularly well at low FPRs (Figure 3).

**Table 2.** Comparison of discriminatory performance of the markers sLR, Re, and Lp using a symbolic regression versus a logistic regression. PPV was based on a 4 % prevalence of PE. Rates given in percentages.

| **Symbolic regression**<br>**QLattice/Feyn v. 2.1.** |  |  | **Logistic regression** |  |  |
| --- | --- | --- | --- | --- | --- |
| **DR (95% CI)** | **FPR (95% CI)** | **PPV %** | **DR (95% CI)** | **FPR (95% CI)** | **PPV %** |
| 40 (31-48) | 4 (1-6) | 30 | 7 (2-11) | 3 (1-5) | 10 |
| 52 (43-60) | 11 (7-15) | 16 | 33 (25-41) | 6 (4-9) | 18 |
| 71 (63-79) | 26 (21-31) | 10 | 65 (57-74) | 25 (20-29) | 10 |

CI: confidence interval; DR: detection rate; FPR: false positive rate; PPV: positive predictive value

**Figure 3.**
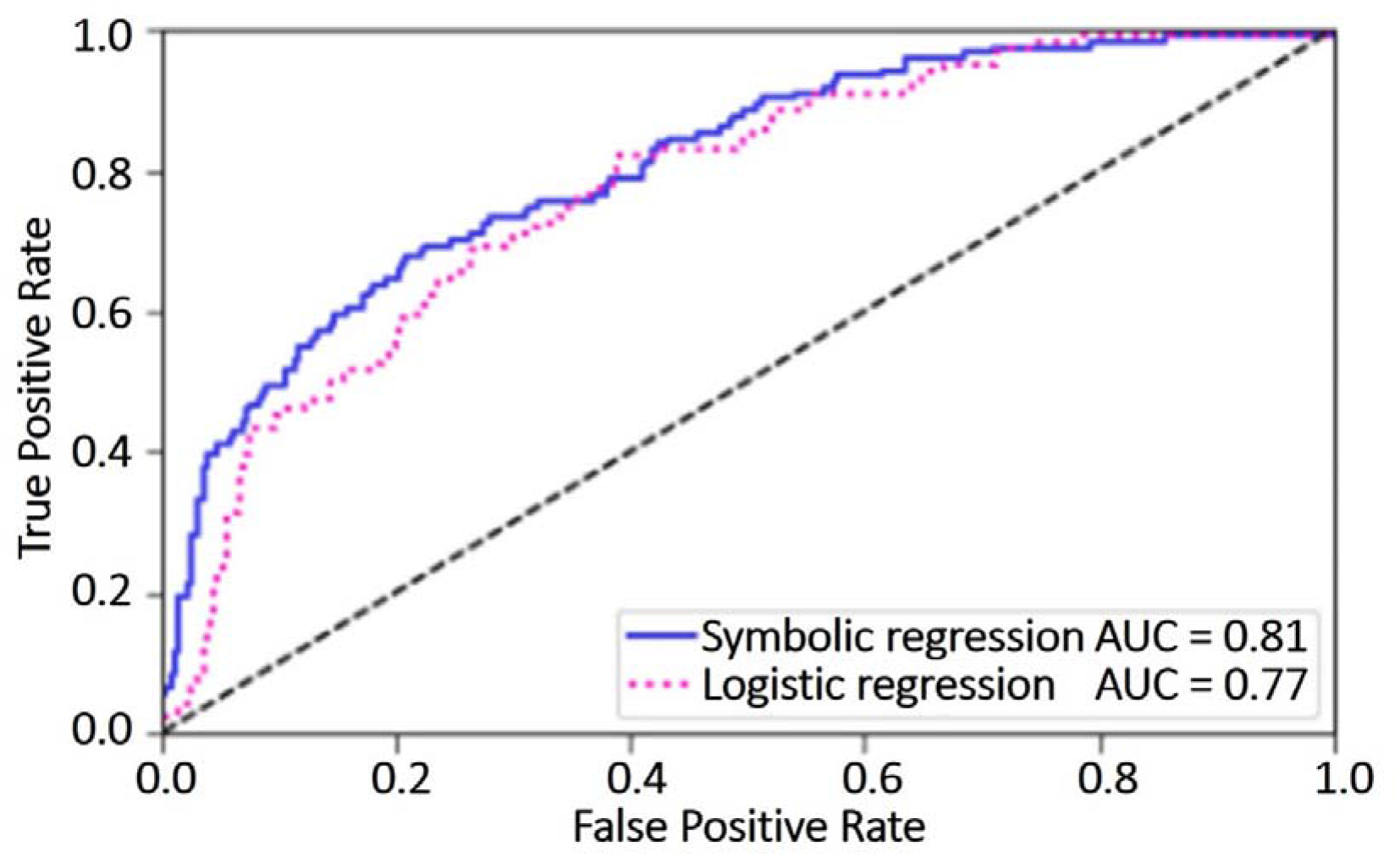
Modelled ROC curves of the discriminatory function using symbolic regression and an optimised discriminatory function defined through logistic regression.

Maternal weight was – despite not being part of the final symbolic regression model – examined as a singular marker, and it was found to be associated (AUC = 0.62) in a Gaussian way with PE (Figure 4).

**Figure 4.**
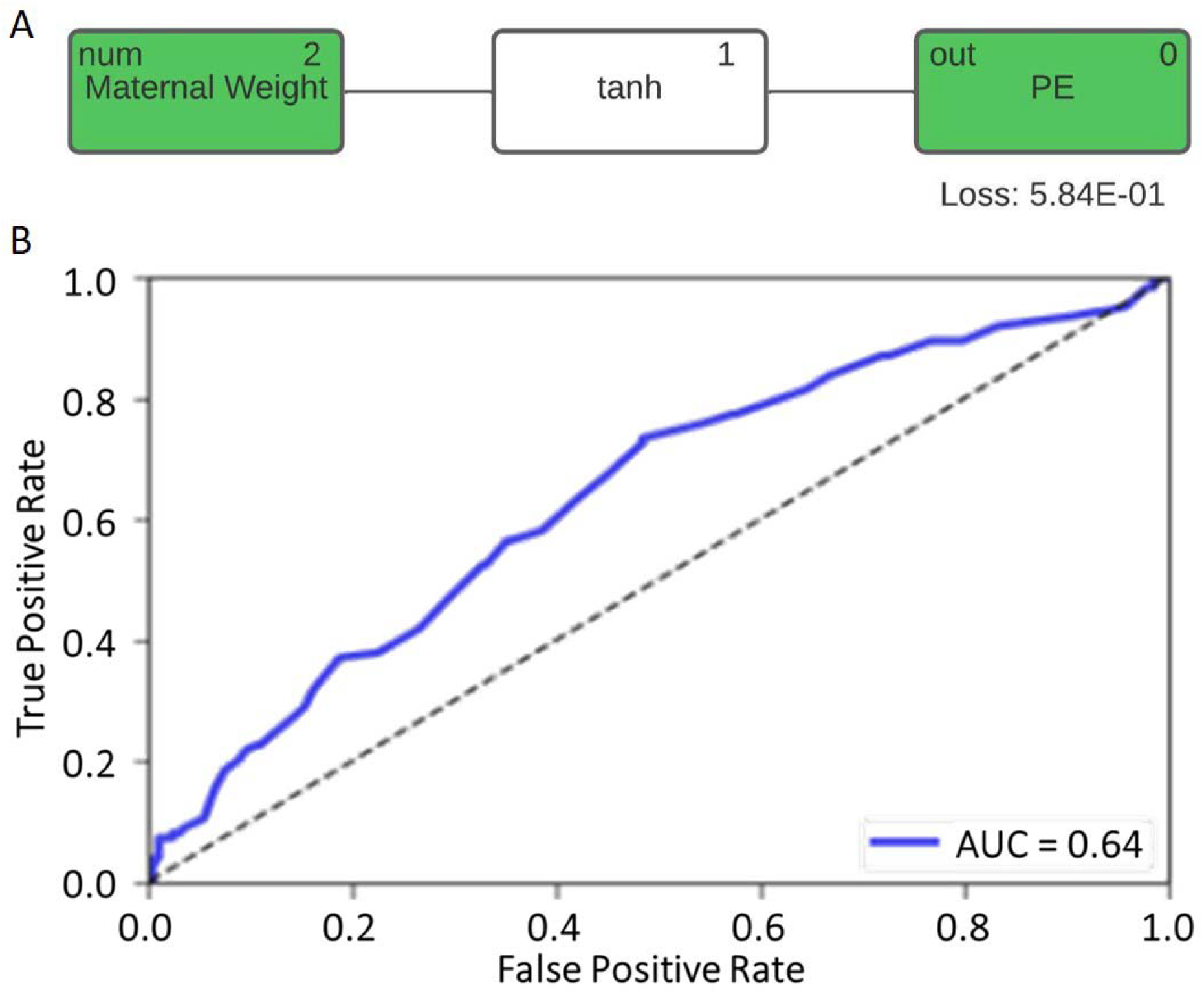
**A**. Model signal path of maternal weight in the symbolic regression model for PE; **B**. The modelled ROC curve with the regressor function of maternal weight as marker for PE.

The three significant contributors to the symbolic regression model, sLR, Lp and Re were studied as singular markers (Figure 5A) and sLR was found to be exponentially related to PE with an AUC of the ROC of 0.76 (Figure 5B); Lp was log-linearly related to PE with an AUC of 0.71 (Figure 5B), whereas Re exhibited a Gaussian relation to PE with an AUC of 0.66 (Figure 5B). For comparison, the same ROC curves were depicted for the logistic regression model of each of the three markers. The AUCs for Lp (AUC = 0.71) and for sLR (AUC = 0.76) were identical to the values obtained in the SR model. This is not surprising, as the relation optimized through the symbolic regression procedure is either a linear or exponential function (closely approximated by the logistic regression). This in contrast to Re, where the logistic regression results in an AUC of 0.56, as compared to the 0.66 in the SR model respectively (Figure 5). The symbolic regression model is thus superior in defining a discriminatory function for Re as compared to the logistic regression model, because it can identify the Gaussian interaction.

**Figure 5.**
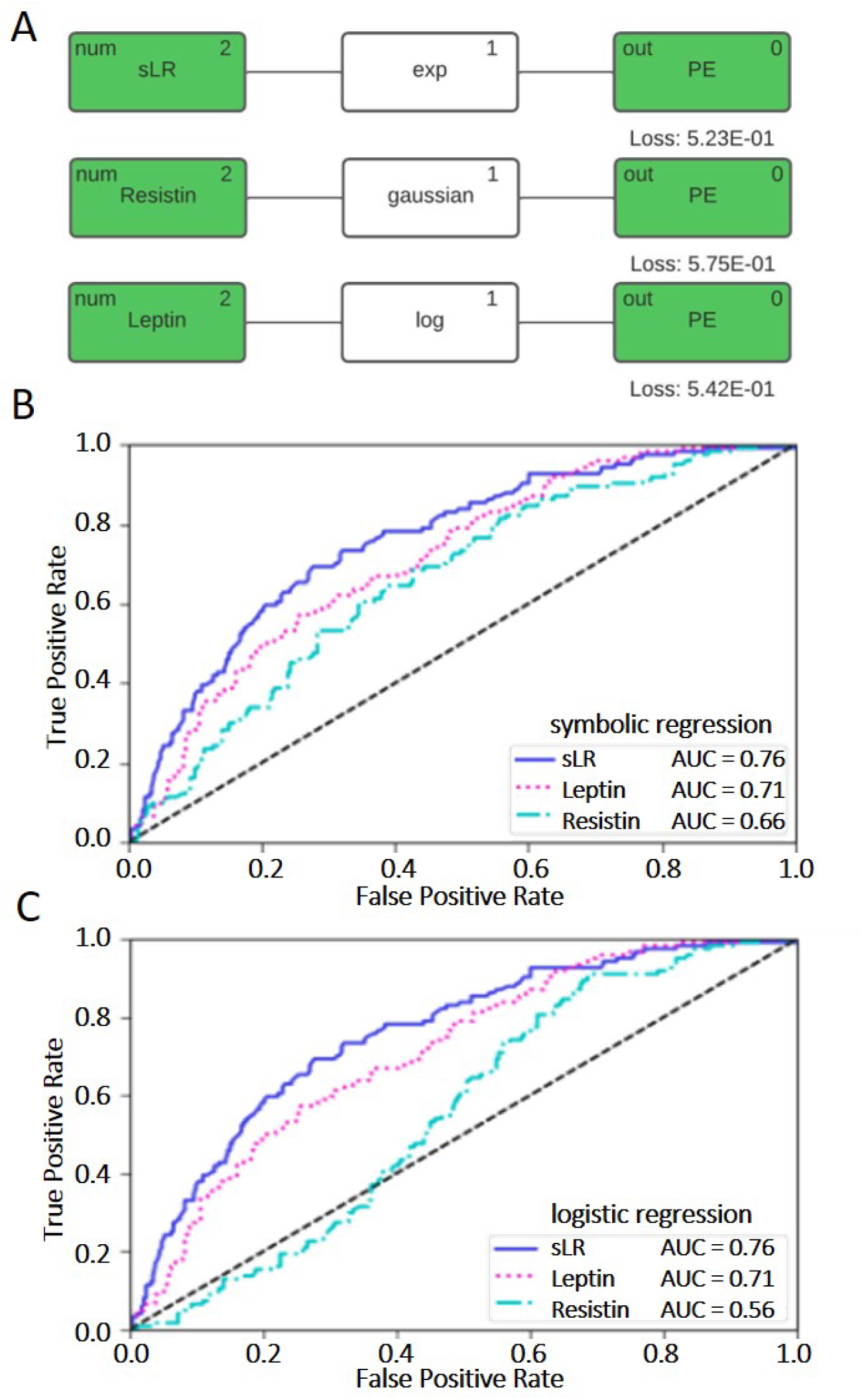
**A**. Model signal paths in the symbolic model for PE for each of the parameters Lp, sLR, and Re individually. **B**. Modelled ROC curves of the symbolic regression models for each of the parameters. **C**. Modelled ROC curves of the logistic regression models for each of the parameters.

## Discussion

The important finding in this study is that symbolic regression using the QLattice/Feyn procedure defines a non-linear relation between Re, Lp, sLR, and PE (Figure 1) that is superior to models based on logistic regression. The AUC increased from 0.77 using the logistic regression model to 0.81 using the symbolic regression model. More importantly, the symbolic regression model performed markedly better for low values of FDR when used as a screening algorithm (Figure 3 and Table 2).The reason for this difference is explained by the ability for the QLattice model to capture the Gaussian interaction between Re and Lp. Previous ML studies of biomarkers in PE have – in very small samples – obtained AUCs of 0.90 – 0.96, including many more markers [51-53]. In this study we have not, except for maternal weight, included maternal risk factors [15], so it must be expected that a clinical performance will be better. Furthermore, the large majority of our cases were non-severe pregnancies giving birth after gestational week 34 (Table 1), and these cases are more difficult to detect with screening markers [14], why one might speculate that the symbolic regression discrimination function would function much better in detecting early and severe pre-eclampsia. However, the real significance of the symbolic regression approach to defining screening algorithms will need to be assessed in prospective studies including other risk factors, i.e. clinical markers, differentiated socio-economical exposures, and health care policies [54].

Overfitting is a risk of all ML approaches, however the QLattice/Feyn2.1 has been shown empirically to exhibit an extent of overfitting comparable to that of linear regression [34], Furthermore, the confidence intervals in Table 1 are of comparable size, reflecting that logistic regression and Qlattice/feyn2.1 symbolic regression do indeed have similar degrees of overfitting. And a level of overfitting much better than other ML approaches [34].

The fact that Re – an inflammatory marker expressed in adipose tissue [24, 55] - is only a marker for PE for specific values of Lp and sLR – suggest that the Re inflammatory function is quenched in extremely low-weight or extremely high weight individuals. This finding may explain the U-shape of the relation between maternal weight and risk of PE [1]. Previous studies have differed in the Re levels measured in maternal serum in PE pregnancies [24, 56, 57], this may – in view of the non-linear interactions described above - possibly be explained by interference from other hormones. The “white-box” nature of symbolic regression may enable a much more detailed understanding of the complicated interplay between different endocrine axes, e.g. the GH – IGF axis [58], the IGF system [59], the placenta-brain-axis [60], in pregnancy. This knowledge may in turn aid in defining new therapeutic targets and screening algorithms.

The demonstration of the potential significance of including non-linear marker interactions should thus lead to a more widespread use of statistical models defined through the “white-box” characteristic of symbolic regression. With the availability of technically and financially sustainable methods to apply symbolic regression to identify non-linear interactions in physiological – and pathophysiological – systems we may well see a revolution in clinical areas where personalized medicine has been very difficult to introduce.

## Highlights

- Capturing non-linear relationships between markers can provide useful insight into physiology and pathophysiology.
- Symbolic regression can be used to produce superior models of the relation between adipokines and PE
- Symbolic regression may help produce more efficient screening algorithms for PE.

## Data Availability

All data produced in the present work are contained in the manuscript

## Acknowledgments

We gratefully acknowledge the expert technical assistance of Pia Lind, and Pernilla Rasmussen. We also gratefully acknowledge the financial support of the Danish Medical Research Council, Copenhagen University, The John and Birthe Meyer Foundation, The Ivan Nielsen Foundation, The Else and Mogens Wedell-Wedellsborg Foundation, The Dagmar Marshall Foundation, The Egmont Foundation, The Fetal Medicine Foundation, The Augustinus Foundation, The Gangsted Foundation, The A.P. Møller Foundation, The Mads Clausens Foundation, The Copenhagen Hospital Corporation, SAFE Network of Excellence and Statens Serum Institut. This research has been conducted using the Danish national Biobank resource, supported by the Novo Nordisk Foundation.

